# Development and Validation of Machine Learning-Based Prediction of Depression Progression Using EHR Data: A Multi-Institutional Retrospective Cohort Study

**DOI:** 10.1101/2025.11.28.25341207

**Authors:** Pegah Ahadian, Angela Fragano, Tianyuan Guan, Qiang Guan, Sophia Z. Shalhout

## Abstract

**Background:** Depression is a leading cause of global disability. Timely identification of patients at risk for clinical worsening remains a major challenge. Electronic health records (EHRs) facilitate large-scale, real-world analyses of disease trajectories. However, standardized symptom scale data such as the Patient Health Questionnaire-9 are often unavailable or recorded only as unstructured text. In this context, International Classification of Diseases (ICD10) diagnostic-code based severity progression provides a pragmatic alternative for developing predictive tools to identify worsening depression.

**Objective:** We aim to develop and evaluate machine-learning and deep-learning models for predicting ICD10-defined progression from mild to moderate/severe depression using EHR data curated by the MedStar Health Research Institute (MHRI).

**Methods:** We conducted a multi-institutional retrospective cohort analysis using the MHRI EHR database, which integrates data from 10 hospitals and 300 outpatient sites across the mid-Atlantic. Adults (≥18 years) with an initial ICD10 diagnosis of mild depression between 2017 and 2023 were included (N=2131). Nonprogressors were defined as patients whose mild major depressive disorder remained mild for 24 months (N=270). Progressors were defined as patients who developed moderate or severe ICD10 depression within 24 months of the index diagnosis (N=533). Data were stratified and split into (60%) training, (20%) validation, and (20%) test subsets. A heterogeneous feature set spanning demographics, healthcare utilization, socioeconomic indices, diagnostic context, and laboratory measurements were available. Logistic regression utilized elastic net regularization with fivefold cross validation, and random forest hyperparameters were tuned by grid search. XGBoost, CatBoost, and a deep neural network (DNN) were trained with standard learning rate, depth, class weighting, and early stopping. A deterministic top model selection framework applied prespecified thresholds of sensitivity at least 0.70 and AUC at least 0.70, and composite rankings integrated accuracy, sensitivity, specificity, and the overfitting gap.

**Results:** The analytic cohort included 803 patients with complete two-year follow-up. Under the selection criteria, the DNN failed to meet the AUC threshold (0.671) and was excluded. Among the remaining models, XGBoost achieved the top composite score (accuracy = 0.72; AUC = 0.776; sensitivity = 0.77; specificity = 0.63; overfit gap = 0.112). Logistic regression ranked second (accuracy = 0.71; AUC = 0.797; sensitivity = 0.79; specificity = 0.61; overfit gap = 0.052), followed by CatBoost and random forest, the latter penalized for overfitting (gap = 0.278). The TinyLlama audit note, generated through a local Hugging Face pipeline, confirmed XGBoost as the most balanced model.

**Conclusions:** Using EHR data from a multi-institutional regional health system, we developed and validated machine-learning models that predicted progression of depression. XGBoost demonstrated the most reliable composite performance. These findings support the feasibility of leveraging socioeconomic and EHR data to predict worsening depression and emphasize the importance of transparent model-selection frameworks for trustworthy clinical artificial intelligence.

## INTRODUCTION

Depression is one of the largest burdens of disease worldwide.^1^ Globally, the prevalence of major depressive disorder (MDD) continues to increase and ranks among the highest causes of disability-adjusted life years (DALYs) and years lived with disability (YLDs).^2^ Unfortunately, a notable acceleration of this trend was observed due to the COVID-19 pandemic, which was associated with a substantial increase in new and persistent depressive episodes worldwide.^3^ The widespread adoption of electronic health records (EHRs) has enabled large-scale, real-world investigations of mental health and clinical trajectories of depression. Multiple systematic reviews highlight the growing utility of artificial intelligence/machine learning (AI/ML) and deep learning (DL) methods applied to EHR data for the diagnosis, risk stratification, and prognosis of depressive disorders.^4,5,6^ In addition, utilizing the International Classification of Diseases (ICD10) depression diagnostic codes in EHRs has demonstrated the feasibility of identifying depression phenotypes and the onset or recurrence of depression.^4,7^ However, several gaps remain. While many studies focus on onset or presence of depression, relatively few have addressed progression of depression defined as the transition from mild to moderate or severe disease states, especially using longitudinal EHR data with ICD10 severity coding.

An important unmet need in mental-health care is the development of predictive models that enable early identification of patients whose depression will progress. Timely intervention by providers can help prevent chronicity, reduce functional disability and suicide risk, and lower health-care utilization and costs.^8,9^ However, many large-scale EHR datasets lack complete standardized depression symptom-scale measures such as the Patient Health Questionnaire-9 (PHQ-9) which can indicate progression of depression. PHQ-9 outcomes are often unstructured, incomplete, unavailable, or inconsistently documented, limiting their use for predictive modeling. To address this limitation, investigators have increasingly turned to ICD diagnostic codes as pragmatic proxies for depression severity and longitudinal progression. Prior studies have demonstrated that ICD-coded diagnosis sequences can approximate symptom-based trajectories and facilitate large-scale phenotyping when structured depression survey data are lacking.^10^ Furthermore, validation studies have shown that ICD10 diagnostic codes can reliably identify depression and track its progression in administrative or EHR data, achieving high specificity (>99%) and positive predictive value (∼90%), thereby providing a robust alternative to symptom-scale instruments like the PHQ-9 when such data are sparse or unavailable.^11^ Together, these findings suggest that ICD-based phenotyping provides a reliable and scalable foundation for developing predictive models of depression progression, particularly in real-world datasets where standardized symptom instruments are incomplete.

In this context, our study leverages data from the MedStar Health Research Institute (MHRI) EHR repository to develop and evaluate machine-learning and deep-learning models for predicting ICD10-defined progression from mild to moderate/severe depression within two years of mild MDD diagnosis. By focusing on severity progression within 24 months in a real-world, multi-institutional EHR cohort and applying a transparent model-selection framework, our work aims to extend the literature on depression prognosis and EHR-driven predictive modeling for early intervention applications.

## METHODS

### Study cohort and outcomes

We performed a retrospective cohort study approved by the Kent State University and Mass General Brigham Institutional Review Boards under a Data Usage Agreement with MHRI. We conducted a multi-institutional analysis using the MHRI EHR database, which integrates longitudinal patient data from ten hospitals and more than 300 affiliated outpatient facilities across the mid-Atlantic region of the United States. The MHRI database includes structured and semi-structured data encompassing demographics, laboratory test values, Social Vulnerability Index (SVI) and Area Deprivation Index (ADI) features, encounter records, ICD10 diagnostic codes, and procedures. Adult patients aged 18 years or older with at least one MDD ICD10 diagnosis recorded between January 1, 2017 and December 31, 2023 were identified (N=47,292) in MHRI. Subjects with an initial qualifying ICD10 diagnosis of mild MDD (F32.0 or F33.0) and at least two depression-related encounters (F32.0, F32.1, F32.2, F32.3, F33.0, F33.1, F33.2, F33.3) within two years of the index mild episode were identified. To ensure sufficient longitudinal follow-up, patients were required to have a minimum of two years of continuous EHR activity following their index diagnosis. A total of 2,131 patients met these initial inclusion criteria.

The target outcomes were defined by ICD10-coded severity depression progression over 24 months. Nonprogressors were defined as patients whose diagnostic trajectory remained limited to mild MDD codes throughout the 24-month observation window, without any recorded moderate, severe, or unspecified MDD codes, nor any diagnostic codes suggestive of bipolar disorder, psychotic depression, or other affective disorders that could confound depression severity classification. Progessors were defined as patients who developed moderate or severe MDD (F32.1, F32.2, F33.1, F33.2, F33.3) within 24 months of the mild index diagnosis. Patients lacking an initial mild MDD diagnosis, and/or continuous follow-up were excluded. The final analytic cohort included 803 patients who met all inclusion criteria. This framework operationalizes ICD10-encoded severity change as a reproducible marker of clinically meaningful worsening.

### Predictor Variables

From the MHRI EHR database, 292 candidate features were extracted. These included demographic, and socioeconomic variables (age, sex, race, ethnicity, insurance, zip code, ADI, SVI), encounter-level utilization metrics (total encounters, encounter type, reason for visit, admission type), laboratory test results (metabolic, hematologic, thyroid, endocrine, inflammatory panels, chemistry, specialty assays) and comorbidity indicators for common chronic conditions (hypertension, diabetes, chronic kidney disease, asthma, chronic obstructive pulmonary disease, heart failure, coronary artery disease, liver disease, arthritis, cancer, epilepsy, multiple sclerosis, Parkinson’s disease, Alzheimer’s disease, and dysthymia). This multi-domain feature offers a comprehensive representation of each patient’s biological, clinical, and contextual risk profile for depression progression.^12^ Continuous missing values were imputed using the mean. Categorical missing values were imputed using the most frequent category. Categorical features were one-hot encoded, and continuous features were standardized using Z-score normalization.

### Feature Selection

Data were partitioned using stratified sampling into 60 percent training, 20 percent validation, and 20 percent test subsets. Feature selection was conducted exclusively within the training subset to avoid information leakage. Interaction-only polynomial feature construction and univariate screening reduced the initial set to the top 100 features for the Logistic Regression (LR) model. A Random Forest (RF) model trained on the training split provided additional feature ranking. Fixed random seeds and the predefined split ensured reproducibility. Stability checks using alternative importance measures showed consistent ranking among the strongest predictors. The final feature set contained 20 predictors.

### Development of AI and ML Models

Five supervised binary classifiers were trained to predict progression including LR, RF, XGBoost, CatBoost, and a DNN. The training split was used for model fitting, and the validation split guided hyperparameter tuning. LR used elastic net regularization with five-fold cross-validation (CV). RF tuning explored tree number, depth, and split and leaf thresholds. XGBoost and CatBoost were trained with fixed learning rate, depth, subsampling, class weighting, and early stopping or calibration on the validation split. The DNN consisted of four fully connected layers with batch normalization, dropout, Adam optimization, and early stopping. Final performance was evaluated exclusively on the held-out test data split.

### Evaluation Metrics and Model Selection

Evaluation metrics included accuracy, area under the receiver operating characteristic curve (AUC), sensitivity, specificity, and F1-score. To prevent overfitting and ensure transparent ‘best’ model selection, a deterministic selection framework was applied, requiring models to meet prespecified performance thresholds of AUC ≥ 0.70 and sensitivity ≥ 0.70. Models that failed to meet these criteria were excluded from final comparison. For models meeting threshold criteria, a composite ranking score was computed. This score integrates accuracy, sensitivity, specificity, and the overfitting gap, defined as the difference in AUC between training and test sets. The ranking approach prioritized models with balanced discrimination and generalization performance. Model interpretability and explainability were further assessed by generating SHapley Additive exPlanations (SHAP) feature importance plots for the top three models and by performing a TinyLlama audit via a local Hugging Face pipeline to confirm reproducibility of results.

### Code and Analysis

All preprocessing and predictive models were implemented in Python 3.13.2 using pandas, NumPy, scikit-learn, TensorFlow, Matplotlib, and Seaborn, with all dependencies managed via pip 25.2. The AI ReAct-style judge and audit note were run in the same Python 3.13.2 environment, using the Hugging Face transformers stack to call TinyLlama/TinyLlama-1.1B-Chat-v1.0 through a CPU-only text-generation pipeline. All preprocessing, model-training, and evaluation steps adhered to the Transparent Reporting of a Multivariable Prediction Model for Individual Prognosis or Diagnosis (TRIPOD+AI) guidelines.^13^

## RESULTS

### Participant Characteristics

A total of 47,292 adult patients with any depression diagnosis were identified in the MHRI EHR database between 2017 and 2023. Of these, 2,172 patients had an initial diagnosis of mild depression. After restricting patients to those with at least two subsequent depression-related encounters recorded within two years of their initial mild episode, 2,131 patients remained eligible for analysis. The study cohort selection process is shown in **Figure 1**. Patients were classified into two outcome groups based on longitudinal ICD10 coding. Cohort 1 (Nonprogessors) were patients whose follow-up codes remained mild throughout the two-year observation window (N = 270). Cohort 2 (Progressors) were patients who progressed to moderate or severe MDD within two years (N = 533) of the index mild depression diagnosis. The final analytic cohort included 803 patients, representing those with feature availability and continuous EHR activity. The final dataset was divided into training (60 %), validation (20 %), and test (20 %) splits using stratified sampling to preserve class proportions. Baseline demographic, and socioeconomic characteristics of each cohort are summarized in **Table 1**. Progressors and nonprogressors were predominantly female, consistent with the known higher prevalence of depression in women.^14^ Both groups showed broadly similar age and race distributions. Across social vulnerability measures (SVI themes and ADI national rank) and healthcare utilization (total encounters), the variables displayed large standard deviations with substantial overlap, indicating that these characteristics were not strongly differentiated between groups on an absolute basis.

**Figure 1:**
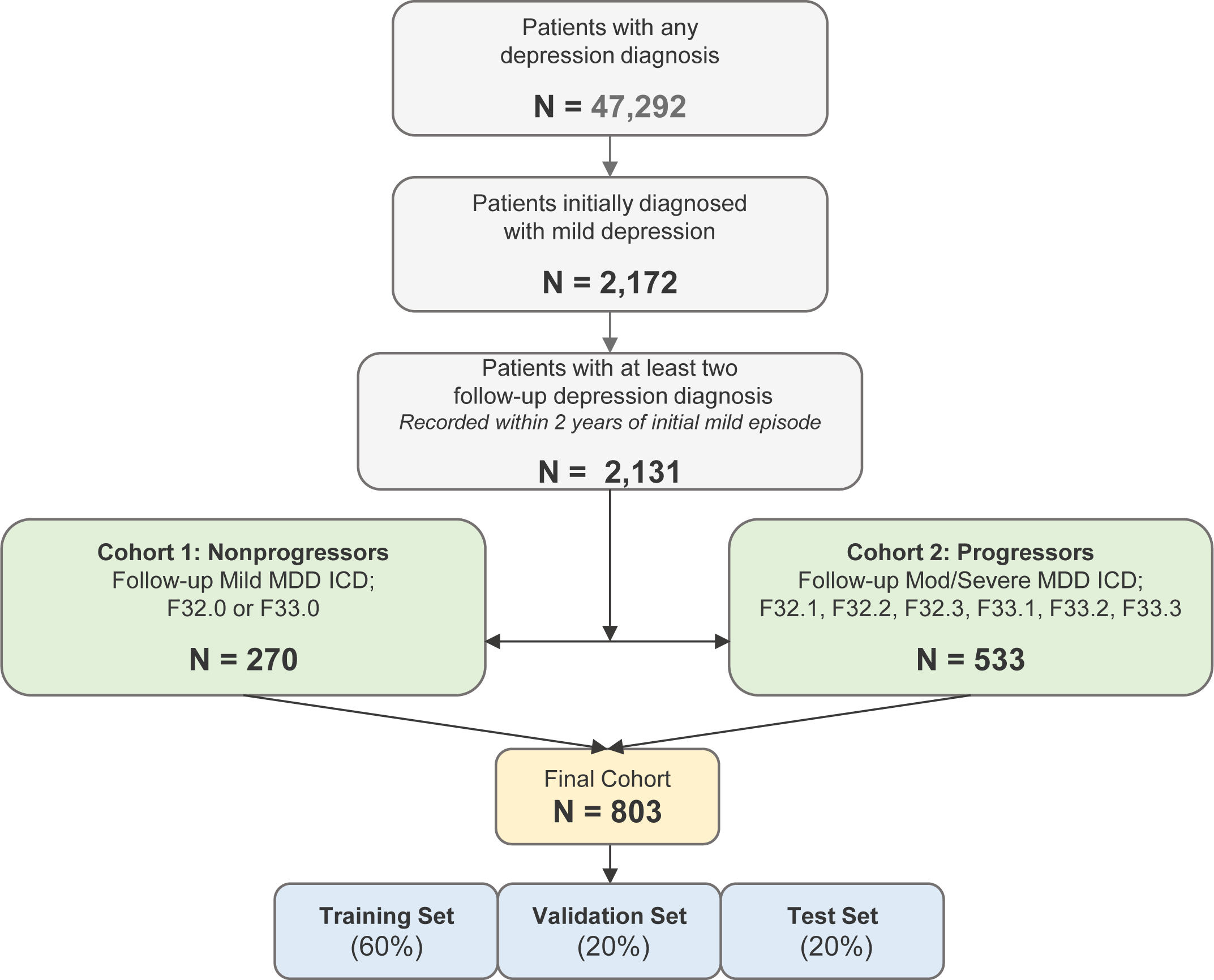
Study Cohorts. Flow diagram illustrating sequential inclusion criteria applied to the MHRI EHR dataset. From 47,292 adults with any depression diagnosis, 2,172 individuals had an initial ICD10 diagnosis of mild MDD. Of these, 2,131 had at least two follow-up depression-related encounters within two years. Patients were classified into ‘Nonprogressors’ (mild-only follow-up codes; N = 270) and ‘Progressors’ (progressed to moderate or severe ICD-10 depression; N = 533). The final study cohort (N = 803) was stratified and divided into training (60 percent), validation (20 percent), and held out test (20 percent) sets for model development and evaluation.

**Table 1:**
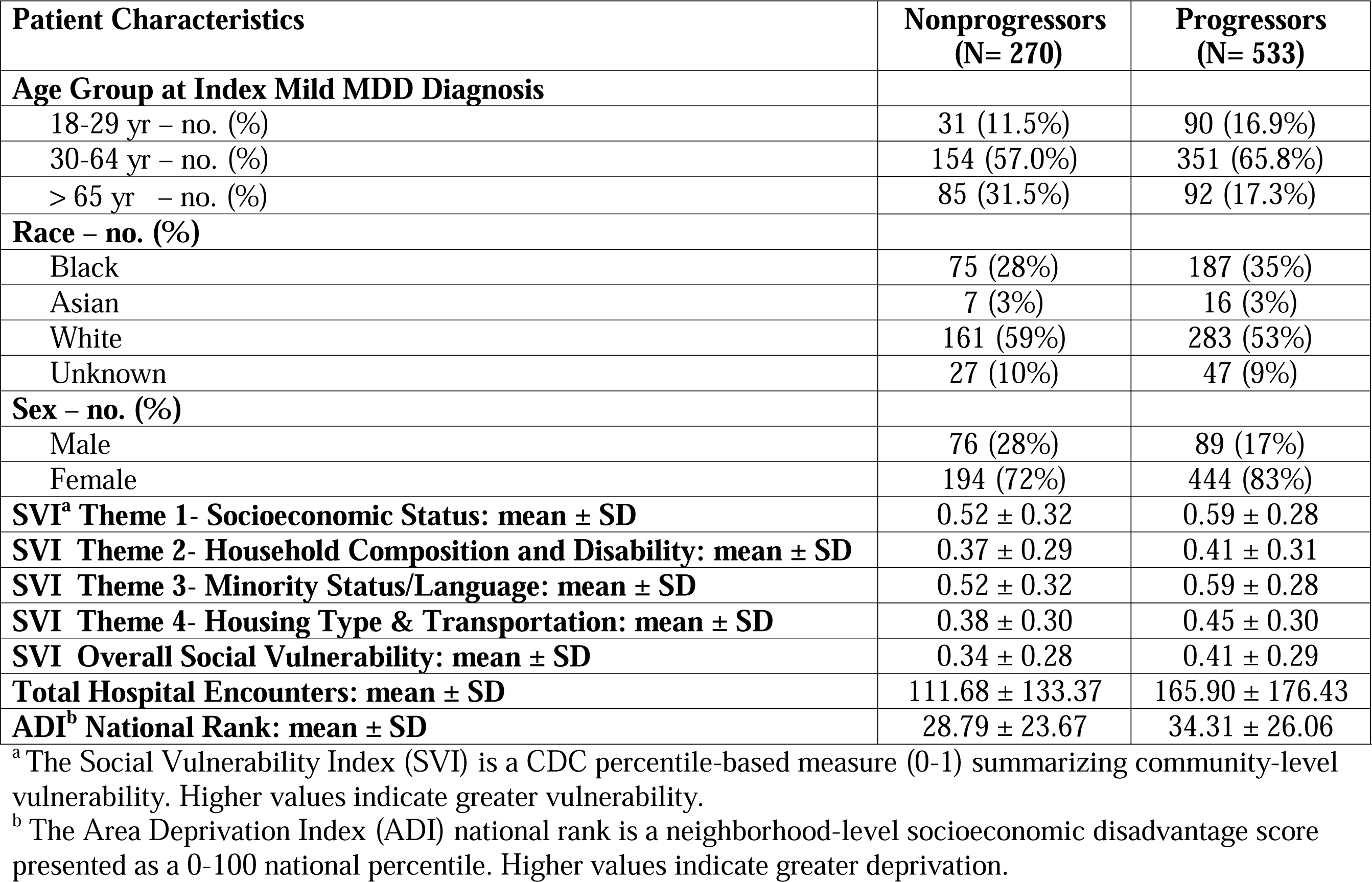
Patient Characteristics.

| **Patient Characteristics** | **Nonprogressors**<br>**(N= 270)** | **Progressors**<br>**(N= 533)** |
| --- | --- | --- |
| **Age Group at Index Mild MDD Diagnosis** |  |  |
| 18-29 yr – no. (%) | 31 (11.5%) | 90 (16.9%) |
| 30-64 yr – no. (%) | 154 (57.0%) | 351 (65.8%) |
| > 65 yr – no. (%) | 85 (31.5%) | 92 (17.3%) |
| **Race – no. (%)** |  |  |
| Black | 75 (28%) | 187 (35%) |
| Asian | 7 (3%) | 16 (3%) |
| White | 161 (59%) | 283 (53%) |
| Unknown | 27 (10%) | 47 (9%) |
| **Sex – no. (%)** |  |  |
| Male | 76 (28%) | 89 (17%) |
| Female | 194 (72%) | 444 (83%) |
| **SVI<sup>a</sup> Theme 1- Socioeconomic Status: mean ± SD** | 0.52 ± 0.32 | 0.59 ± 0.28 |
| **SVI Theme 2- Household Composition and Disability: mean ± SD** | 0.37 ± 0.29 | 0.41 ± 0.31 |
| **SVI Theme 3- Minority Status/Language: mean ± SD** | 0.52 ± 0.32 | 0.59 ± 0.28 |
| **SVI Theme 4- Housing Type & Transportation: mean ± SD** | 0.38 ± 0.30 | 0.45 ± 0.30 |
| **SVI Overall Social Vulnerability: mean ± SD** | 0.34 ± 0.28 | 0.41 ± 0.29 |
| **Total Hospital Encounters: mean ± SD** | 111.68 ± 133.37 | 165.90 ± 176.43 |
| **ADI<sup>b</sup> National Rank: mean ± SD** | 28.79 ± 23.67 | 34.31 ± 26.06 |

<sup>a</sup> The Social Vulnerability Index (SVI) is a CDC percentile-based measure (0-1) summarizing community-level vulnerability. Higher values indicate greater vulnerability.
<sup>b</sup> The Area Deprivation Index (ADI) national rank is a neighborhood-level socioeconomic disadvantage score presented as a 0-100 national percentile. Higher values indicate greater deprivation.

### Model Training and Evaluation

The LR, RF, XGBoost, CatBoost, and DNN were trained on selected features. Hyperparameter tuning on the validation split yielded stable convergence. Early stopping was used for XGBoost, CatBoost, and the DNN, while LR and RF hyperparameters were optimized using cross-validation. Deterministic preprocessing, fixed random seeds, and a predefined data split were employed to ensure reproducibility. Models were evaluated against prespecified performance thresholds of AUC ≥ 0.70 and sensitivity ≥ 0.70. The evaluator loads the metrics per-split, applies our chosen policy on test sensitivity and AUC, rejects models that are considered ‘violators’ with explicit reasons, then scores the remaining models using a weighted composite of AUC, F1-weighted, specificity, accuracy, and an overfit penalty based on the train-test AUC gap. Finally, models are sorted and ranked by total score, emitting a ranked report. In addition, a brief rationale contrasting the models ranked as #1 and #2 is provided. The large language model (LLM) produces a concise audit note; its prompt contains the fixed policy and the top-k results with exact metrics, with a brief explanation of why the model ranked first is chosen and how the model ranked second compares.

Under these settings, the DNN did not meet the AUC requirement (AUC = 0.671 on the test set) and was excluded from further comparison. The remaining LR, CatBoost, XGBoost and RF models exceeded threshold criteria and were retained for composite ranking. Test-set performance metrics are summarized in **Table 2**. XGBoost demonstrated the most balanced performance (accuracy = 0.72, AUC = 0.78, sensitivity = 0.77, specificity = 0.63) and the train, validation, and test AUC curves for the XGBoost model (**Figure 2A**) illustrate moderate overfitting consistent with a train-test AUC gap of 0.11. The corresponding test-set confusion matrix (**Figure 2B**) demonstrates moderate sensitivity and specificity. Logistic regression achieved the highest AUC (0.80) with a sensitivity = 0.79. AUC curves for training, validation and test show minimal divergence, consistent with the small overfitting gap (0.05) (**Figure 3A**). The test-set confusion matrix for the LR model (**Figure 3B**) illustrates balanced classification performance across the two outcome classes. CatBoost demonstrated effective control of overfitting via ordered boosting and early stopping (**Figure 4A**) with similar accuracy (0.71) and sensitivity (0.72) (**Figure 4B**). Random forest produced high sensitivity (0.85) but lower specificity and a larger overfitting gap (0.28), reducing its composite ranking. XGBoost received the highest composite score (TOTAL = 70.36), followed by LR (TOTAL = 65.85), CatBoost (TOTAL = 65.73), and RF (TOTAL = 56.00). The audit note generated by the pipeline (**Figure 5**) summarized the trade-offs between the top models, highlighting the stronger F1 and accuracy of XGBoost relative to logistic regression, and the sensitivity-specificity balance that distinguished the three surviving boosted or linear models. These rankings confirmed XGBoost as the top-scoring model and provided a concise explanation for its selection over the logistic regression model based on differences in weighted-F1, accuracy, and specificity profiles.

**Figure 2:**
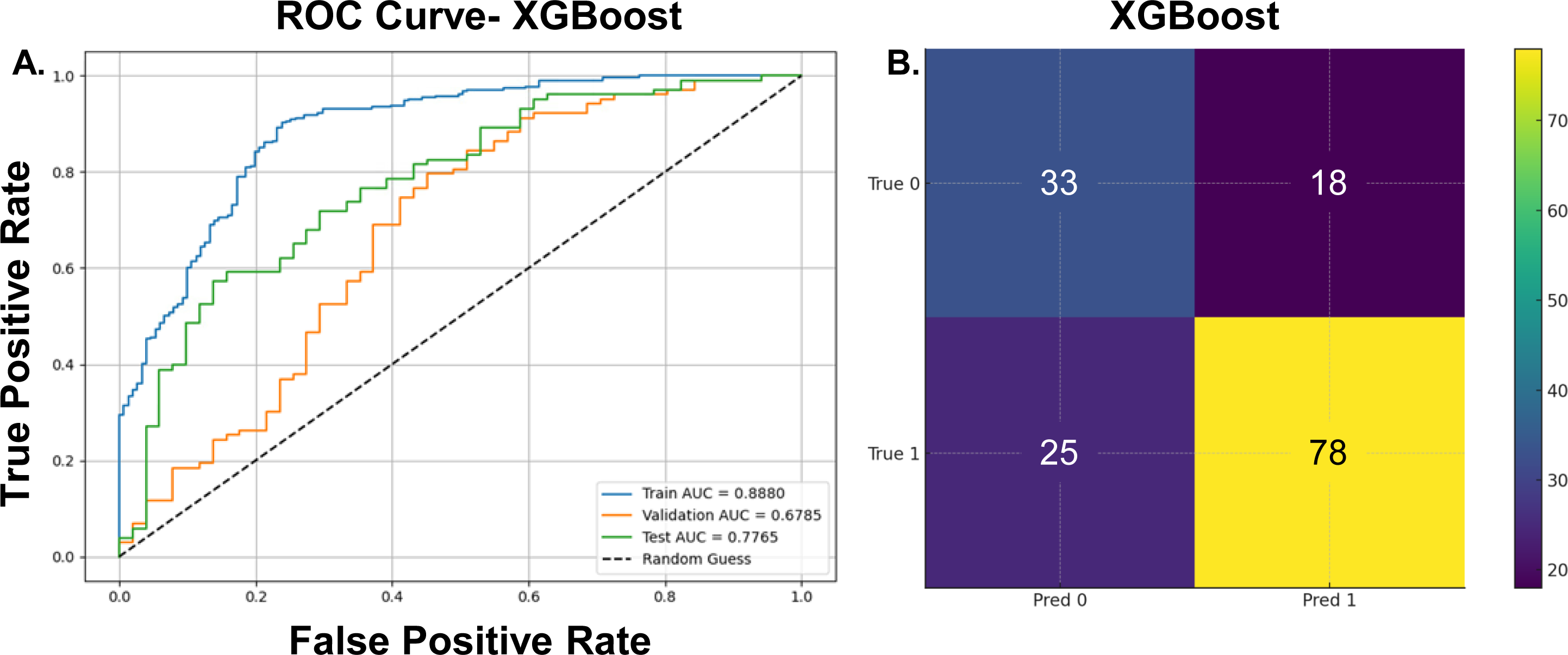
XGBoost Model Performance. A. Receiver operating characteristic curves for the XGBoost classifier evaluated on the training, validation, and test splits. B. Test-set confusion matrix for XGBoost is depicted. The model correctly identified 33 true negatives and 78 true positives, with 18 false positives and 25 false negatives, reflecting the model’s observed test-set sensitivity (0.77) and specificity (0.63).

**Figure 3:**
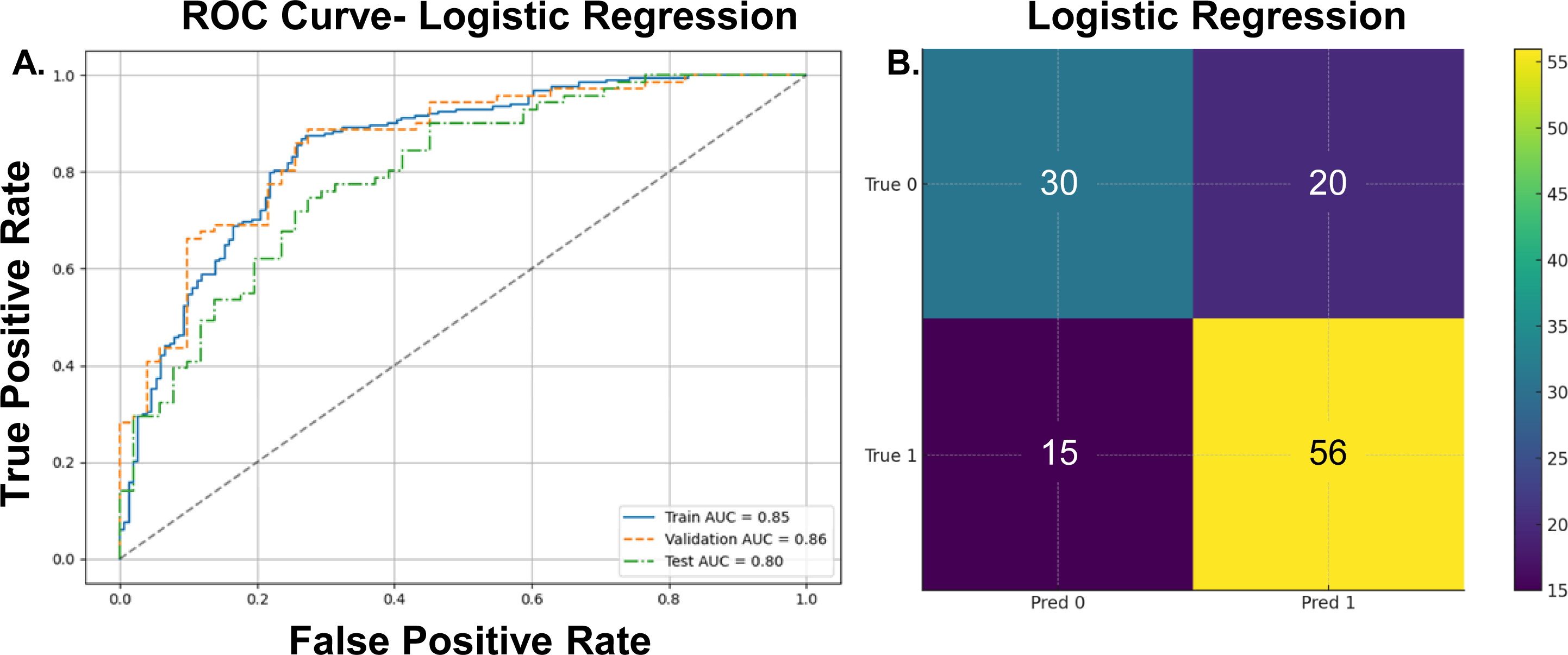
Logistic Regression Model Performance. A. Receiver operating characteristic curves for the LR classifier evaluated on the training, validation, and test splits. The model achieved AUC = 0.85 on the training set, AUC = 0.86 on the validation set, and AUC = 0.80 on the independent test set, indicating stable generalization across splits. B. Test-set confusion matrix for LR model is shown.

**Figure 4:**
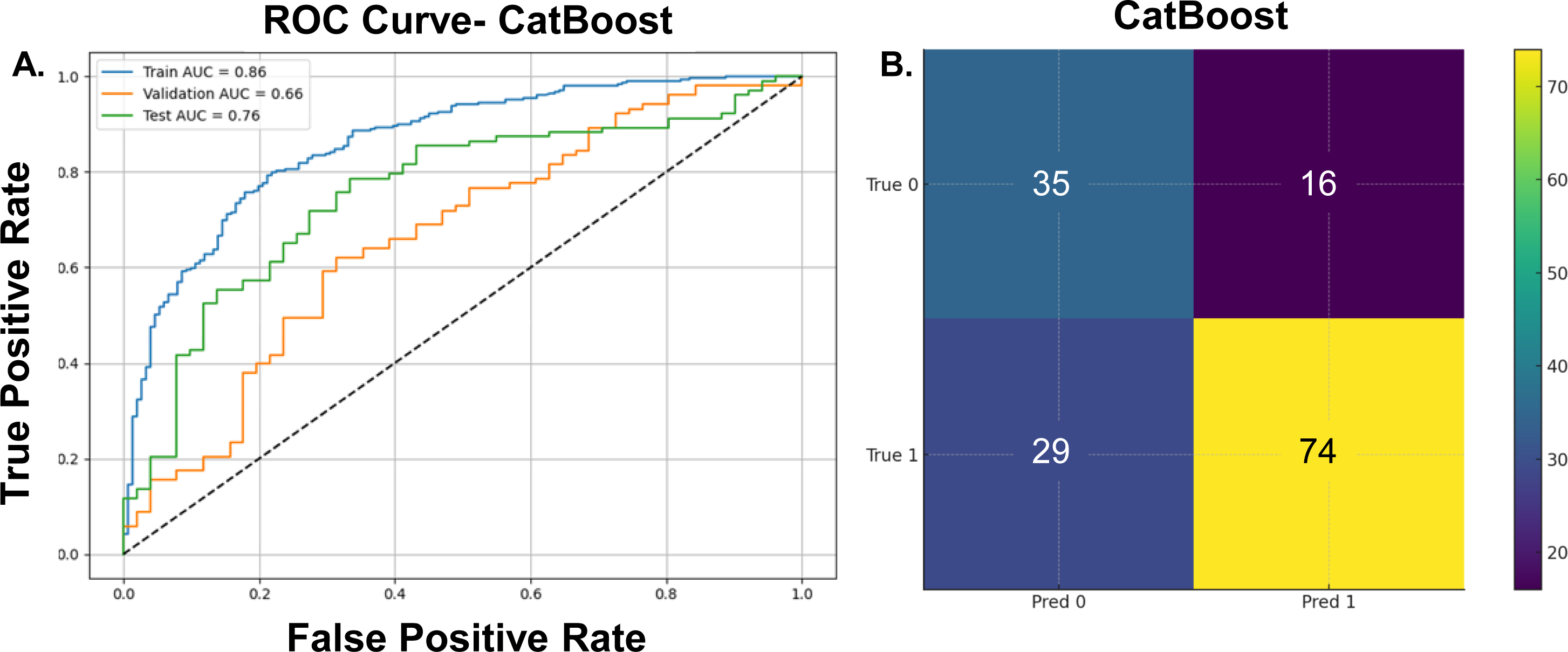
CatBoost Model Performance. A. The ROC curves display CatBoost performance across the training, validation, and test sets. The model achieved AUC = 0.86 on the training set, AUC = 0.66 on the validation set, and AUC = 0.76 on the test set, reflecting moderate generalization. B. The CatBoost classifier produced 35 true negatives, 16 false positives, 29 false negatives, and 74 true positives on the held-out test set. These results illustrate a balanced recall profile.

**Figure 5:**
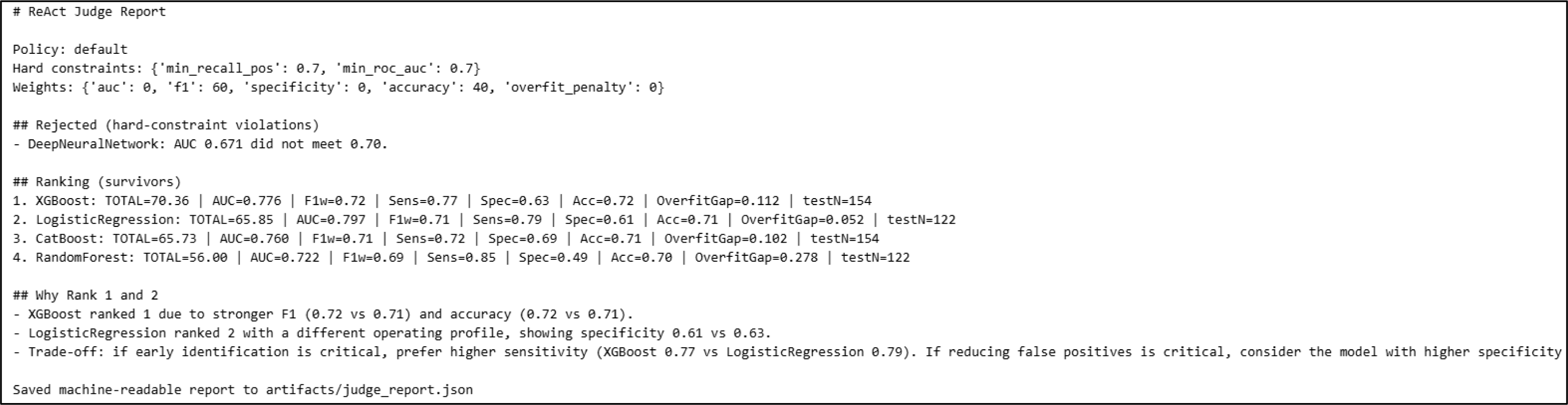
Model Ranking and ReAct Audit Output. This figure presents the full output of the ReAct-style model evaluation and auditing pipeline used to rank classifiers. The top block lists the enforced hard constraints for model acceptance, including minimum sensitivity (recall) and minimum ROC-AUC. The middle section displays the ranked models that satisfied these constraints, with each model’s composite score, AUC, F1-weighted score, sensitivity, specificity, accuracy, overfitting gap, and test set size. The bottom section summarizes the audit rationale comparing the top-ranked models, explaining why XGBoost was selected as the preferred model and how logistic regression performed in comparison.

**Table 2:** Summary of Model Performance on Test Data.

| **Model** | **Accuracy** | **F1-weighted** | **AUC** | **Sensitivity** | **Specificity** | **Overfitting Gap** | **Model Status** |
| --- | --- | --- | --- | --- | --- | --- | --- |
| XGBoost | 0.72 | 0.72 | 0.776 | 0.77 | 0.63 | 0.11 | Retained |
| LR | 0.71 | 0.71 | 0.797 | 0.79 | 0.61 | 0.05 | Retained |
| CatBoost | 0.71 | 0.71 | 0.760 | 0.72 | 0.69 | 0.10 | Retained |
| RF | 0.70 | 0.69 | 0.722 | 0.85 | 0.49 | 0.28 | Retained |
| DNN | 0.64 | 0.64 | 0.671 | - | - | - | *Rejected* |

### Model Explainability

The XGBoost SHAP summary plot (**Figure 6A**) highlights ‘Total Encounters’, ‘Age at Visit Group’, and ‘Number of Social Categories’ exerting the largest influence on predicted risk of progression. Several SVI-related features (RPL themes) demonstrate modest but directionally consistent effects, indicating that social vulnerability contributes incremental predictive signal within the model’s multivariate context. The SHAP distribution for logistic regression (**Figure 6B**) highlights ‘Age at Visit Group’, ‘Total Encounters’, ‘Immature Platelet Fraction’, and ‘Total Bilirubin’ as the strongest contributors to model output. The CatBoost SHAP summary plot (**Figure 6C**) identifies ‘Total Encounters’, ‘Diagnostic Priority’, and SVI-related variables (RPL Theme 1, RPL Theme 3, RPL Theme 4) as the most influential predictors. CatBoost produced more compact SHAP ranges than XGBoost, consistent with its smoother boosting procedure.

**Figure 6:**
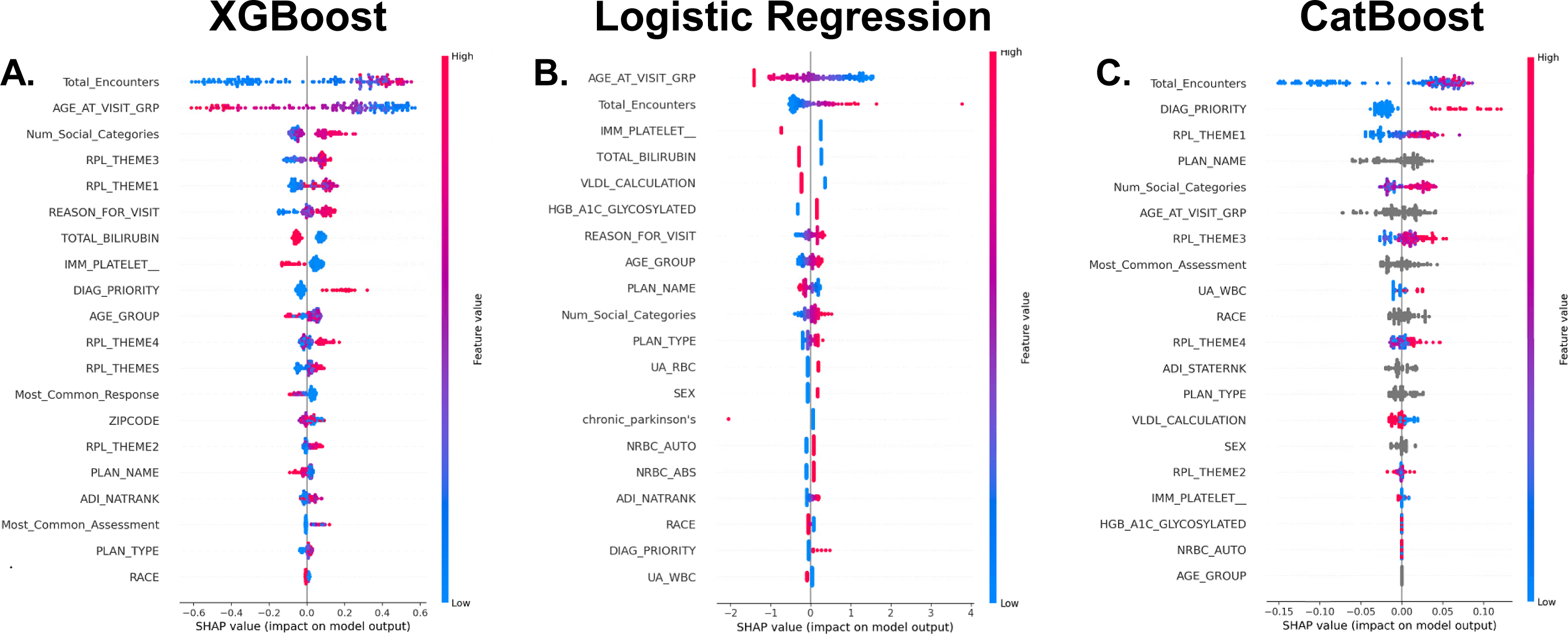
SHAP Feature Importance Across Top Models. (A) XGBoost SHAP summary plot which depicts ‘Total encounters’ and ‘Age group at visit’ with the largest positive contributions to predicted progression, and multiple SVI-related variables (RPL_THEME1, RPL_THEME3) exerting smaller but consistent effects. Higher feature values (pink) generally increase predicted risk, while lower values (blue) reduce it. (B) Logistic Regression SHAP summary plot depicting ‘Age group at visit’ and ‘Total encounters’ as the dominate factors influencing the model. (C) CatBoost SHAP summary plot is shown. Predictions are driven by a blend of encounter frequency, diagnostic priority, RPL themes, and laboratory values. CatBoost displays tighter SHAP distributions, reflecting stronger regularization and monotonic handling of categorical features.

## DISCUSSION

We evaluated machine-learning and deep-learning models to predict ICD10-defined progression from mild to moderate or severe MDD using real-world EHR data. Depression is a heterogeneous disorder influenced by biological, social, and healthcare-access factors. Early prediction and identification of high-risk patients remain a major clinical priority.^15^ Our classification framework operationalized severity changes strictly through ICD10 codes, enabling reproducible labeling within large EHR systems despite the well-known absence or inconsistency of symptom-scale data such as PHQ-9.^16–18^ Using a curated cohort of 803 adults, we demonstrated that structured socioeconomic and EHR data can provide meaningful predictive signal for clinically relevant worsening of depression.

XGBoost achieved the most balanced performance across discrimination, calibration stability, and sensitivity-specificity trade-offs compared to the remaining models. Logistic regression achieved the highest AUC, likely retaining competitive performance due to strong and well-selected predictors. SHAP-based explainability analyses highlighted heterogeneous risk signatures distributed across laboratory markers, healthcare-utilization patterns, chronic comorbidities, and socioeconomic vulnerability indices. This aligns with our current understanding of depression as a multi-system disorder influenced by several factors.^12^ Notably, SVI and ADI measures contributed non-trivially to model predictions, consistent with prior studies linking such socioeconomic factors with depression.^19–20^ However, the large standard deviations and overlapping distributions across groups underscore that no single factor is sufficient to discriminate progression risk, at least in this cohort, reinforcing the need for multivariable modeling (**Table 1**).

Several important limitations must be emphasized. First, as previously mentioned, the dataset did not include standardized symptom scales such as PHQ-9, which limits granularity in measuring depressive severity. ICD10 codes provide an imperfect proxy for symptom-level severity, since coding decisions may vary by clinician, healthcare setting, and reimbursement policies. ICD10 misclassification bias, for example, may attenuate observed associations or obscure true predictive signal. Second, although the cohort was multi-institutional, the final analytic sample (N=803) reflects only patients with continuous two-year follow-up, potentially introducing selection bias toward patients with higher healthcare utilization. Third, EHR-derived features are influenced by patterns of care rather than purely underlying pathophysiology, and some predictors may reflect differential access or utilization rather than intrinsic risk such as ‘total hospital encounters’ and ‘insurance type’. In addition, a larger, external, multicenter cohort is required to test our models’ robustness and confirm that the performance observed, generalize across populations and EHR systems. While trained and evaluated on a multi-institutional cohort, MHRI is a single regional health system, which may limit external generalizability, particularly given geographic, socioeconomic, and practice-pattern differences across regions.

We applied strict data-splitting and leakage controls. However, overfitting remains possible, as reflected by the overfitting gaps observed in our tree-based models. Finally, the study period overlapped with the COVID-19 pandemic, which introduced major disruptions to healthcare utilization, diagnostic coding patterns, and follow-up frequency. These system-level changes may have influenced both the exposure features, such as encounter counts, and the progression labels derived from ICD10 codes. As a result, some observed lack of progressions may partially reflect pandemic-related care patterns rather than true underlying clinical trajectory. Future models should assess performance in post-pandemic cohorts.^21–23^

Despite these constraints, the study demonstrates that gradient-boosted tree approaches applied to socioeconomic and EHR data can identify individuals at risk for clinically coded worsening of depression with reasonable accuracy. The methodological framework, stratified splitting, leakage-free feature selection, calibration checks, standardized model auditing, and composite model ranking, aligns with recommended practices for clinical machine-learning transparency and reproducibility.^24^ Future work should incorporate symptom-scale data, natural-language-processing extraction of narrative clinical assessments, and causal-inference frameworks to distinguish true clinical deterioration from healthcare-utilization artifacts. External validation across other health systems and post-pandemic cohorts will be essential for assessing transportability. Ultimately, integrating interpretable risk models into clinical workflows may support proactive monitoring for targeted intervention aimed at patients most likely to experience worsening depressive illness.

## CONCLUSIONS

In this multi-institutional retrospective cohort study, we demonstrated that structured socioeconomic and EHR data can be used to predict ICD10-defined progression from mild to moderate or severe depression with meaningful accuracy. After standardized preprocessing, transparent data-splitting, and deterministic model selection, XGBoost achieved the strongest overall balance of discrimination, sensitivity, specificity, and generalization, while logistic regression provided the highest AUC and CatBoost offered competitive performance with a distinct operating profile. These findings show that depression-progression risk can be predicted from real-world clinical, laboratory, and socioeconomic features. Our results highlight the feasibility of building reproducible, interpretable machine-learning pipelines for mental-health risk stratification using EHR data. Although further validation in external health systems and prospective settings is needed, the approach provides a scalable framework for future predictive tools that could support earlier identification of patients at risk for worsening depressive illness.

## AUTHORS’ CONTRIBUTIONS

Conceptualization: SZS, PA

Data curation: SZS, PA

Formal analysis: SZS, PA, AF, TG, QG

Funding acquisition: SZS, PA, QG

Investigation: SZS, PA

Methodology: SZS, PA

Project administration: SZS

Resources: SZS, PA

Supervision: SZS, QG

Validation: SZS, PA

Visualization: SZS, PA, AF

Writing-original draft: SZS, AF, PA

Writing-review and editing: SZS, AF, PA, TG, QG

## ABBREVIATIONS

ADI: Area Deprivation Index
AI: Artificial Intelligence
AUC: Area Under the Receiver Operating Characteristic Curve
CPU: Central Processing Unit
CV: Cross-Validation
DALY: Disability-Adjusted Life Year
DNN: Deep Neural Network
EHR: Electronic Health Record
ICD10: International Classification of Diseases, Tenth Revision
LLM: Large Language Model
LR: Logistic Regression
MDD: Major Depressive Disorder
MHRI: MedStar Health Research Institute
ML: Machine Learning
PHQ9: Patient Health Questionnaire-9
RF: Random Forest
ROC: Receiver Operating Characteristic Curve
SVI: Social Vulnerability Index
TRIPOD+AI: Transparent Reporting of a Multivariable Prediction Model for Individual Prognosis or Diagnosis (Artificial Intelligence extension)
YLD: Years Lived With Disability

## Acknowledgements

This work was supported by the AIM-AHEAD Coordinating Center, funded by the NIH. The data used in this study were obtained from the MedStar Health Research Institute (MHRI) electronic health record (EHR) data warehouse. MedStar Health System includes an extensive network of clinical facilities in the mid-Atlantic region, including 10 hospitals (33% rural hospitals) and over 300 points-of-care connected by MedStar’s EHR system, built on the Cerner Millennium platform. The authors thank the MHRI teams for data curation, accession, and training.

## Data Availability

The MedStar Health Research Institute electronic health record data used in this study cannot be publicly shared due to institutional policy and patient-privacy regulations. Access to this data requires a formal Data Use Agreement with MedStar Health and approval from the relevant Institutional Review Boards. Researchers interested in accessing the underlying data should contact the MedStar Health Research Institute Research Administration. All analysis code and model scripts used in this study are available via GitHub.

